# An open resource for T cell phenotype changes in COVID-19 identifies IL-10-producing regulatory T cells as characteristic of severe cases

**DOI:** 10.1101/2020.05.31.20112979

**Authors:** Julika Neumann, Teresa Prezzemolo, Lore Vanderbeke, Carlos P. Roca, Margaux Gerbaux, Silke Janssens, Mathijs Willemsen, Oliver Burton, Pierre Van Mol, Yannick Van Herck, CONTAGIOUS co-authors, Joost Wauters, Els Wauters, Adrian Liston, Stephanie Humblet-Baron

## Abstract

The pandemic spread of the novel coronavirus SARS-CoV-2 is due, in part, to the immunological properties of the host-viral interaction. The clinical presentation varies greatly from individual to individual, with asymptomatic carriers, mild to moderate-presenting patients and severely affected patients. Variation in immune response to SARS-CoV-2 may underlie this clinical variation. Using a high dimensional systems immunology platform, we have analyzed the peripheral blood compartment of 6 healthy individuals, 23 mild-to-moderate COVID-19 patients and 20 severe COVID-19 patients. We identify distinct immunological signatures in the peripheral blood of the mild-to-moderate and severe COVID-19 patients, including T cell lymphopenia, more consistent with peripheral hypo-than hyper-immune activation. Unique to the severe COVID-19 cases was a large increase in the proportion of IL-10-secreting regulatory T cells, a lineage known to possess anti-inflammatory properties in the lung. Annotated data is openly available (https://flowrepository.ors/experiments/2713) with clinical correlates, as a systems immunology resource for the COVID-19 research community.

## Introduction

The novel coronavirus SARS-CoV-2 has infected millions of people worldwide, and impacted the society and the global economy in an unprecedented manner. The pandemic spread is due, in part, to the immunological properties of the host-viral interaction. Clinical presentation of SARS-CoV-2 infections (COVID-19) varies greatly from individual to individual, with asymptomatic carriers, mild-to-moderate-presenting patients and severely to critically affected patients ^1,2,3,4,5,6^. This clinical heterogeneity is responsible for the severity of the pandemic, with asymptomatic cases and long latency increasing the R0, and severe pathology at the other end of the clinical spectrum result in high mortality ^1,2,3,4,5,6,7,8^. Variation in immune response to SARS-CoV-2 may underlie this clinical disparity and unravelling the immunological features associated with progression into severe and life-threatening disease is highly needed in order to guide therapeutic decision making and biomarker discovery.

Immunological investigations during the SARS-CoV-2 pandemic have focused on identifying altered immune responses in COVID-19 patients. Innate immune response is an essential first line defense against viruses, including type I and III interferon (IFN). Recent studies have suggested that SARS-CoV-2 inhibits type I IFN production and signaling ^9^, potentially explaining the long pre-symptomatic period and persistent viral load in many patients. Defects in NK cell function may also be present, with reports of higher expression of activation and exhaustion markers on NK cells, and impaired NK cytotoxicity and cytokine production ^9,10^, and additional problems with innate immunity are likely to be identified. Other studies suggest a defect in the adaptive immune system. Lymphopenia is widely reported in COVID-19, and the severity of lymphopenia has been correlated with disease severity ^10,11,12,13,14,15,16,17,18,19^ Functional defects within T cells have been reported, with an increased number of non-functional CD4^+^ T cells and impaired T cell cytokine production ^10,11,20^. In a series of 38 patients, an increase in naïve CD4^+^ was reported in COVID-19 patients, suggestive of impaired activation ^21^. This was confirmed in an independent study, but without significant changes between moderate and severe patients ^22^, while another study reported no change ^9^ Parallel findings were reported in broncho-alveolar lavage fluid of COVID-19 patients, with enrichment of naïve CD4^+^ T cells in severely affected patients ^18^. Overall, it remains to be determined whether defects in innate or adaptive immunity or a synergistic effect of both underlie the unusually long infectious period of SARS-CoV-2.Immune analysis is also contributing to an understanding of COVID-19 pathology. Parallels with other respiratory infections, such as influenza ^23^, have led to the hypothesis that pathology is immune-mediated rather than due to direct viral induction, and the potential success of immune-modulating therapeutics in small-scale clinical trials provides preliminary support for this model ^24,25,26,27^ Neutrophils seem to be consistently elevated in severe patients and are associated with poor outcomes ^10,11,12,13,14,15,16,17,22,28^. Several studies have identified COVID-19 as a hyper-inflammatory status, with a “cytokine storm” of pro-inflammatory cytokines ^9,11,13,14,18,29,30,31,32,33^ IL-6, in particular, was consistently higher in severely affected patients compared to moderate and milder cases, suggesting it might be associated with disease severity ^11,13,14,18,30,32,33^. Inconsistent findings have been reported regarding the changes in myeloid subsets ^9,12,34,35^ however, severe COVID-19 patients seem to have an increased number of inflammatory monocytes, producing higher levels of IL-6 and GM-CSF ^12,35^. Bronchoalveolar lavage fluid of severe COVID-19 cases also revealed high levels of monocytes and neutrophils as well as a pro-inflammatory environment ^18,36^. Excessive T cell activation has also been suggested as a possible driver of disease, with increased expression of activation markers (such as HLA-DR, CD38, CD69, CD25, CD44, Ki-67, OX40 and CD137) by CD4^+^ and CD8^+^ T cells in severe patients ^9,11,12,20,21,33^ As with the cause of poor viral clearance, the cause of excessive immune pathology remains unclear.

The ambiguity of the COVID-19 immune profile is based, in part, on the recent origin of the pandemic, but also on the design of COVID-19 clinical trials, many of which have substandard design, lacking suitable controls or data transparency ^37^ This probably is especially acute in studies on the phenotypic and functional changes of T cells, where the limited and sometimes contradictory reports may be attributed to the vagaries of cytometry. Altered gating hierarchy, use of alternative markers, different thresholds for defined expression – each can modify the outcome of analysis ^38^. A potential solution is to provide open-access high dimensional data, allowing independent analysis with alternative strategies, while still preserving the rapid publication process required to deal with an evolving pandemic. As part of the CONTAGIOUS consortium, multi-omic approaches are being performed in a systematic and coordinated manner against a longitudinal COVID-19 cohort. Here we provide the first high dimensional flow cytometry analysis for the CONTAGIOUS cohort. The peripheral blood of 6 healthy individuals, 23 mild-to-moderate COVID-19 patients and 20 severe COVID-19 patients, was assessed for changes to the T cell compartment, and identified a signature of IL-10-producing regulatory T cells in those patients with severe COVID-19. Annotated data is openly available (https://flowrepository.org/experiments/2713), allowing transparent and evolving data analysis.

## Results

As part of the CONTAGIOUS study into the immunome of COVID-19, patients were recruited through the University Hospital in Leuven, Belgium, beginning 27th March 2020. The dataset was generated on 6 healthy individuals, 23 mild-to-moderate COVID-19 patients (WHO score 3-4) and 20 severe COVID-19 patients (WHO score 5-7). Demographic data for the patients are summarized in **Table 1**, with individual data on demographic and clinical values in **Supplementary Resource 1**.

**Table 1.** Clinical features of the COVID-19 cohort. Median (25-75 percentiles) for key demographic and clinical parameters of patients entered into the study. BMI, body mass index; CRP, C-reactive protein; n/a, not available; WBC, white blood cell count; WHO, World Health Organization.

| **Parameter** | **Healthy** | **Mild-moderate COVID-19** | **Severe COVID-19** |
| --- | --- | --- | --- |
| Number of patients | 6 | 23 | 20 |
| Age (years) | 56 (40-62) | 58 (53-69) | 64.5 (53-69) |
| Male:female ratio | 3:3 | 10:13 | 14:6 |
| WHO score | n/a | Score 3: 11/ Score 4: 12 | Score 5: 12/ Score 6: 8 |
| BMI | n/a | 25.1 (22.7-31.7) | 29.1 (24.4-32.5) |
| CRP (mg/L) | n/a | 72.35 (27.6-138.9) | 141.2 (74.75-239.2) |
| WBC (10<sup>3</sup>/μL) | n/a | 6.32 (4.07-9.4) | 8.42 (6.72-10.06) |
| Neutrophil count (x10<sup>3</sup>/μL) | n/a | 4.5 (2.9-7.1) | 6.05 (5.23-8.2) |
| Lymphocyte count (x10<sup>3</sup>/μL) | n/a | 1.0 (0.8-1.7) | 0.9 (0.625-1.225) |

In order to determine the T cell phenotypes associated with clinical heterogeneity in COVID-19, we stimulated T cells *ex vivo* and used high parameter flow cytometry covering a comprehensive set of subset markers, activation markers and cytokines. First, we assessed the number of major leukocyte subsets in PBMCs, using key lineage markers (CD3, CD4, CD8, FOXP3, CD14, CD19). Cluster-based flow cytometry analysis, pooling all samples, separated leukocytes into populations that corresponded to CD4^+^ T cells, CD8^+^ T cells, CD19^+^ B cells, CD14^+^ monocytes and CD4^-^CD8^-^ T cells (**Figure 1A,B, Supplementary Figure 1**). Traditional gating was used to quantify these populations, and FOXP3^+^ regulatory T cells, in each sample. Despite the marked lymphopenia in COVID-19 patients (**Figure 1C**), the relative proportion of these leukocyte populations remained unchanged with COVID-19 patients (**Figure 1D**), other than a trend towards lower CD8 T cell numbers, indicating a non-specific mechanism of leukopenia. Subtle, but significant, differences were observed in the expression of lineage markers, with comparison of pooled mild-moderate and severe COVID-19 patient samples showing a more similar expression pattern than pooled healthy controls (**Figure 1E**). At the individual level, healthy individuals were largely clustered together, while mild-moderate and severe COVID-19 patients exhibited similar cellular phenotypes (**Figure 1F,G**). Together, these data indicate only subtle shifts in the balance of blood leukocyte populations with COVID-19, without any discrimination between mild-moderate and severe cases.

**Figure 1.**
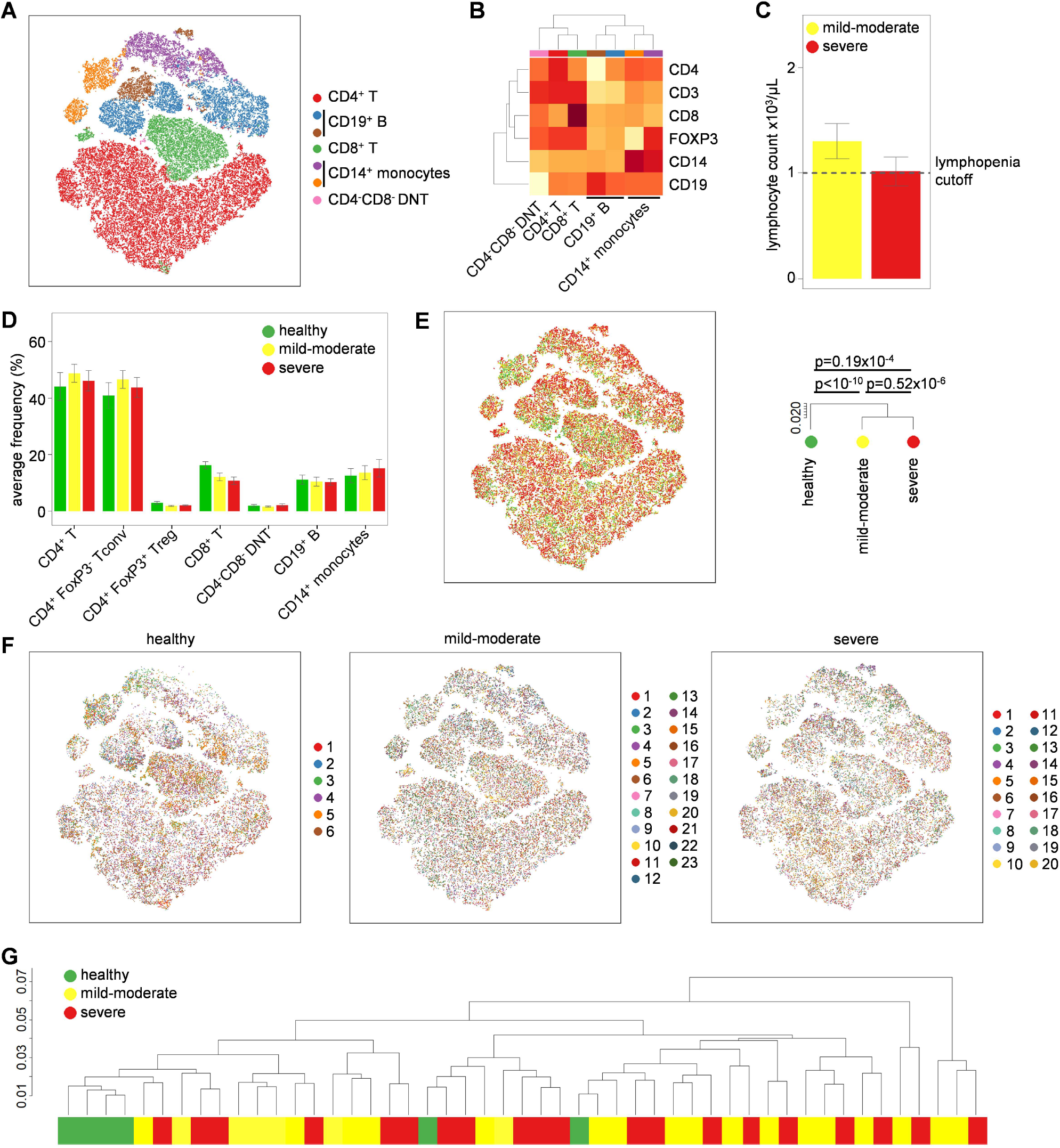
Peripheral blood leukocyte changes in mild-moderate and severe COVID-19 patients. PBMCs were isolated from healthy controls (n=6), mild-moderate COVID-19 patients (n=23) and severe COVID-19 patients (n=20), stimulated and assessed through flow cytometry. **(A)** tSNE representation of PBMC populations based on the expression of lineage markers: CD3, CD4, CD8, FOXP3, CD19, CD14. FlowSOM clusters were annotated based on **(B)** expression of lineage markers across each cluster (see also Fig. S1). **(C)** Clinical lymphocyte counts for mild-moderate and severe COVID-19 patients. **(D)** Quantification of leukocyte subsets in healthy controls, mild-moderate COVID-19 and severe COVID-19 patients based on manual gating. **(E)** tSNE representation of PBMC populations based on aforementioned lineage markers for each condition, namely healthy controls, mild-moderate COVID-19 and severe COVID-19 patients. The dendrogram shows the comparative similarity based on the Kolmogorov-Smirnov statistic calculated using the cross-entropy distributions derived from tSNE. **(F)** tSNE representation of PBMC populations based on aforementioned lineage markers for each condition, namely healthy controls, mild-moderate COVID-19 and severe COVID-19 patients, with each individual represented in a different color. **(G)** Dendrogram showing the comparative similarity between individuals based on the Kolmogorov-Smirnov statistic calculated using the cross-entropy distributions derived from tSNE. Mean ± SEM.

We next investigated the activation and polarization of conventional CD4^+^ T cells in COVID-19 patients. Using the expression of CD45RA, CCR7, 4-1BB, CD25, CTLA-4, HLA-DR, IFN*γ*, IL-2, IL-4, IL-6, IL-10, IL-17a, PD-1, RORγt, T-BET and TNFα, we clustered the conventional CD4^+^ T cell population into 15 biologically-distinct subsets: naïve CTLA-4^-^ cells, naïve CTLA- 4^+^ cells, TCM, TCM CTLA-4^+^, TCM IL-2^+^, TEM, TEM Th1 CTLA4^+^, TEM Th1 CTLA-4^-^, TEM IL-2^+^, TEM TNFα^+^, TEM TNFα^+^ CTLA-4^+^, TEM PD-1^high^, TEMRA, TEMRA Th1 and TEMRA Th17 (**Figure 2A,B, Supplementary Figure 2**). Quantification of these subsets across healthy volunteers and COVID-19 patients demonstrated no significant changes, with only a trend towards increased TCM and decreased TEM being present in severe COVID-19 patients, with mild-moderate patients intermediate in number between healthy and severe COVID-19 patients (**Figure 2C**). Global phenotypic analysis, by contrast, found the cellular phenotype of conventional CD4^+^ T cells to be more similar between healthy individuals and severe COVID-19 patients (**Figure 2D**). At an individual level, the cellular phenotypes of healthy, mild-moderate COVID-19 and severe COVID-19 samples were intermingled (**Figure 2E,F**). This data does not support a peripheral CD4 T cell hyper-activation model of COVID-19.

**Figure 2.**
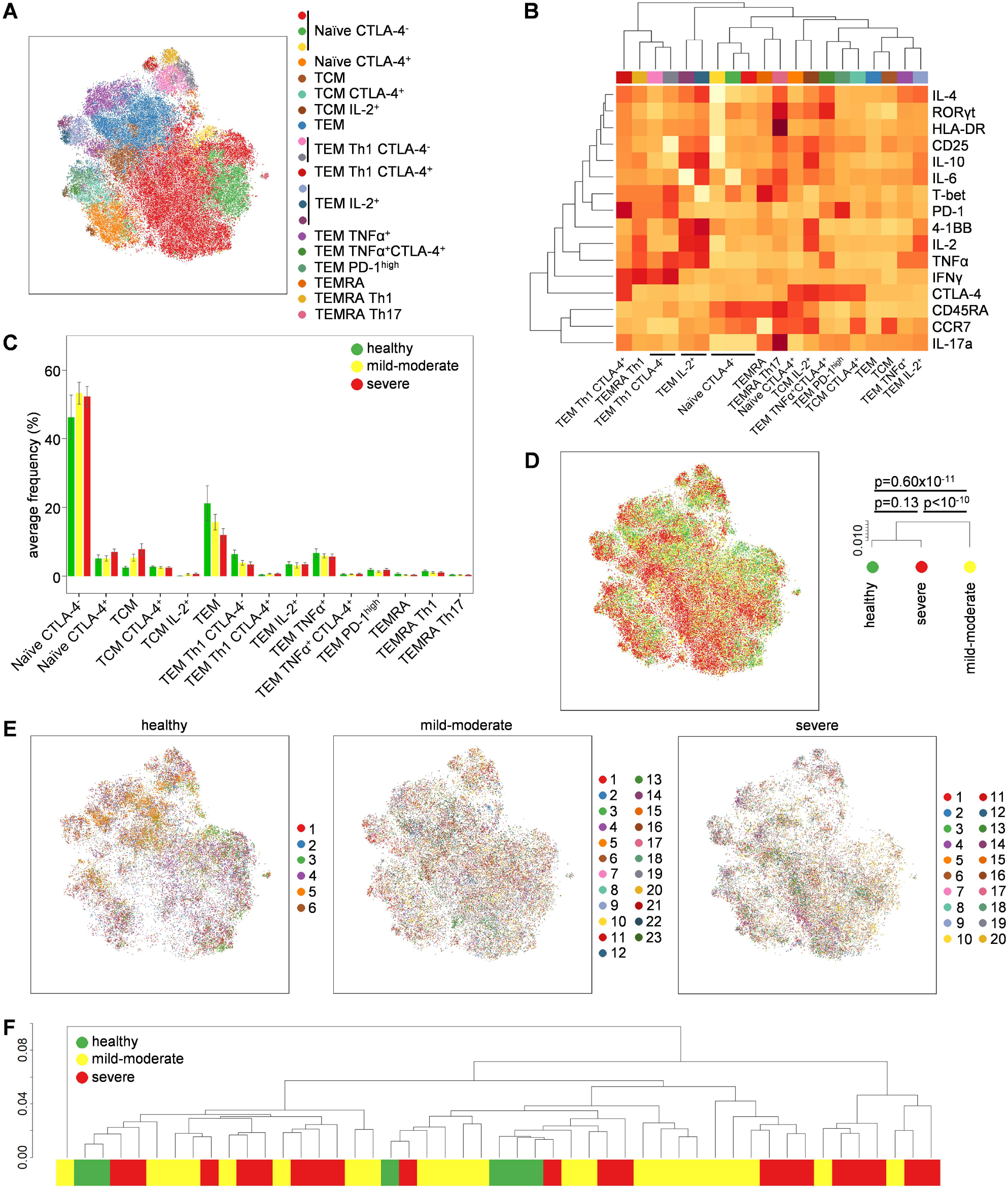
Altered conventional T cell phenotypes in COVID-19. PBMCs were isolated from healthy controls (n=6), mild-moderate COVID-19 patients (n=23) and severe COVID-19 patients (n=20), stimulated and assessed through flow cytometry. CD3^+^CD14^−^CD4^+^CD8^−^FOXP3^−^ conventional T cells were manually gated in FlowJo. **(A)** tSNE representation of conventional CD4 T cell cluster populations based on the expression of phenotypic markers: CD45RA, CCR7, 4-1BB, CD25, CTLA-4, HLA-DR, IFNγ, IL-2, IL-4, IL-6, IL-10, IL-17a, PD-1, RORγt, T-BET, TNFα. FlowSOM clusters were annotated based on **(B)** expression of phenotypic markers across each cluster (see also Fig. S2). **(C)** Quantification of conventional T cell subsets in healthy controls, mild-moderate COVID-19 and severe COVID-19 patients. **(D)** tSNE representation of PBMC populations based on aforementioned phenotypic markers for each condition, namely healthy controls, mild-moderate COVID-19 and severe COVID-19 patients. The dendrogram shows the comparative similarity based on the Kolmogorov-Smirnov statistic calculated using the cross-entropy distributions derived from tSNE. **(E)** tSNE representation of PBMC populations based on aforementioned phenotypic markers for each condition, namely healthy controls, mild-moderate COVID-19 and severe COVID-19 patients, with each individual represented in a different color. **(F)** Dendrogram showing the comparative similarity between individuals based on the Kolmogorov-Smirnov statistic calculated using the cross-entropy distributions derived from tSNE. Mean ± SEM.

Using the same approach, we investigated the phenotype of FOXP3^+^ regulatory T cells. Clustering based on expression markers identified 13 regulatory T cell subsets with biologically distinct characteristics: naïve CTLA-4^-^, naïve CTLA-4^+^, TEM, TEM CTLA-4^+^, TCM, TCM CTLA-4^+^, IL-2 producing CTLA-4^+^, IL-10-producing, TNFα-producing CTLA-4^+^, TNFα-producing CTLA-4^-^, IFNγ-producing CTLA-4^-^, IFNγ-producing CTLA-4^+^ and HLA-DR^+^ cells (**Figure 3A,B, Supplementary Figure 3**). Quantification of these subsets across healthy volunteers and COVID-19 patients demonstrated an increase in TCM CTLA-4^+^ regulatory T cells and IL-10-producing regulatory T cells in severe COVID-19 patients, with mild-moderate patients being intermediate (**Figure 3C**). Manual gating of IL-10-producing regulatory T cells confirmed a 5-fold increase in severe COVID-19 patients only, a significant rise over both healthy individuals and mild-moderate COVID-19 patients (**Figure 3D**). Cellular phenotypes were largely similar across healthy individuals and COVID-19 patients at both the pool (**Figure 3E**) and individual (**Figure 3F,G**) level. While preliminary, the increase in IL-10-producing regulatory T cells in severe COVID-19 patients is of particular interest, given the role of the murine equivalent in controlling lung inflammation ^39^

**Figure 3.**
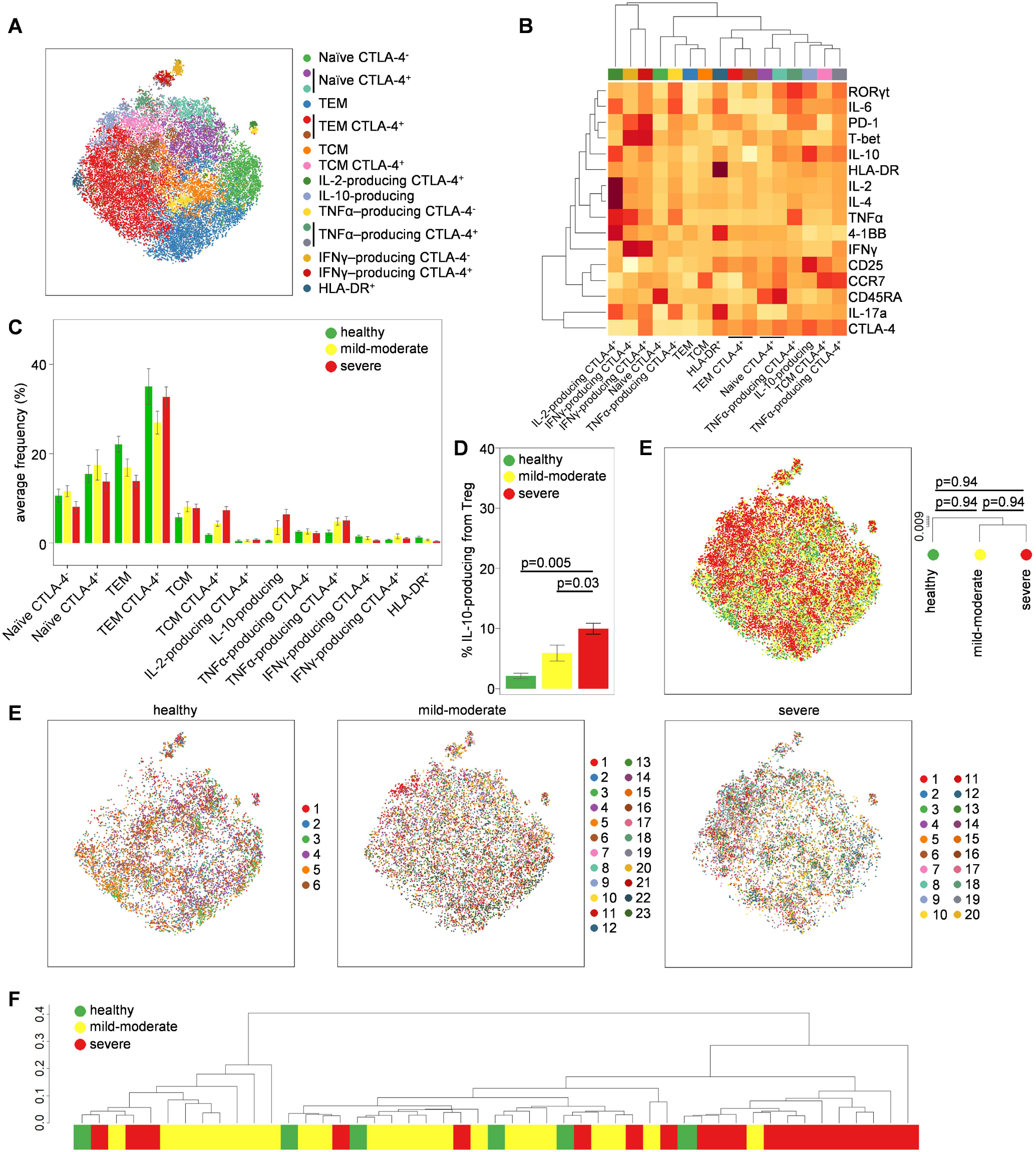
Altered regulatory T cell phenotypes in severe and moderate COVID-19. PBMCs were isolated from healthy controls (n=6), mild-moderate COVID-19 patients (n=23) and severe COVID-19 patients (n=20), stimulated and assessed through flow cytometry. CD3^+^CD14^-^ CD4^+^CD8^-^FOXP3^+^ regulatory T cells were manually gated in FlowJo. **(A)** tSNE representation of regulatory T cells based on the expression of phenotypic markers: CD45RA, CCR7, 4-1BB, CD25, CTLA-4, HLA-DR, IFNγ, IL-2, IL-4, IL-6, IL-10, IL-17a, PD-1, RORγt, T-BET, TNFα. FlowSOM clusters were annotated based on **(B)** expression of phenotypic markers across each cluster (see also Fig. S3). **(C)** Quantification of regulatory T cell subsets in healthy, mild-moderate COVID-19 and severe COVID-19 patients. **(D)** Manually gated IL-10^+^ regulatory T cells in healthy, mild-moderate COVID-19 and severe COVID-19 patients. **(E)** tSNE representation of PBMC populations based on aforementioned phenotypic markers for each condition, namely healthy controls, mild-moderate COVID-19 and severe COVID-19 patients. The dendrogram shows the comparative similarity based on the Kolmogorov-Smirnov statistic calculated using the cross-entropy distributions derived from tSNE. **(F)** tSNE representation of PBMC populations based on aforementioned phenotypic markers for each condition, namely healthy controls, mild-moderate COVID-19 and severe COVID-19 patients, with each individual represented in a different color. **(G)** Dendrogram showing the comparative similarity between individuals based on the Kolmogorov-Smirnov statistic calculated using the cross-entropy distributions derived from tSNE. Mean ± SEM.

Finally, we investigated changes in the CD8^+^ T cell compartment. 12 subsets of CD8^+^ T cells were identified that correlate to biologically-distinct functions: naïve CTLA-4^-^, naïve CTLA-4^+^, TEM, TEM Th1, TEM Tc1 IL-2^+^, TEM Tc1/17, TEM Tc17, TEM IL-2^+^, TEM TNFα^+^, TCM CTLA-4^+^, TEMRA and TEMRA Tc1 (**Figure 4A,B, Supplementary Figure 4**). Quantification of individual subsets did not identify any significant changes (**Figure 4C**), however the highly inflammatory TEM Tc1/Tc17 population (expressing both IFNγ and IL-17) was 13-fold more numerous in COVID-19 patients than in healthy controls, with 27/43 (63%) COVID-19 patients exhibiting numbers >2 standard deviations above the average of healthy individuals (**Supplementary Resource 1**). This population did not discriminate between mild-moderate and severe COVID-19 patients. At a global level, the cellular phenotype of CD8^+^ T cells was largely unchanged by COVID-19-status, using both pooled (**Figure 4D**) and individual (**Figure 4E,F**) analysis. Together this data suggests an ongoing CD8 T cell response in COVID-19 patients, with expansion of the potent Tc1/Tc17 TEM subset, however the comparative weakness of the response, and its failure to predict disease severity, argue against systemic CD8 T cell responses being a pathogenic driver.

**Figure 4.**
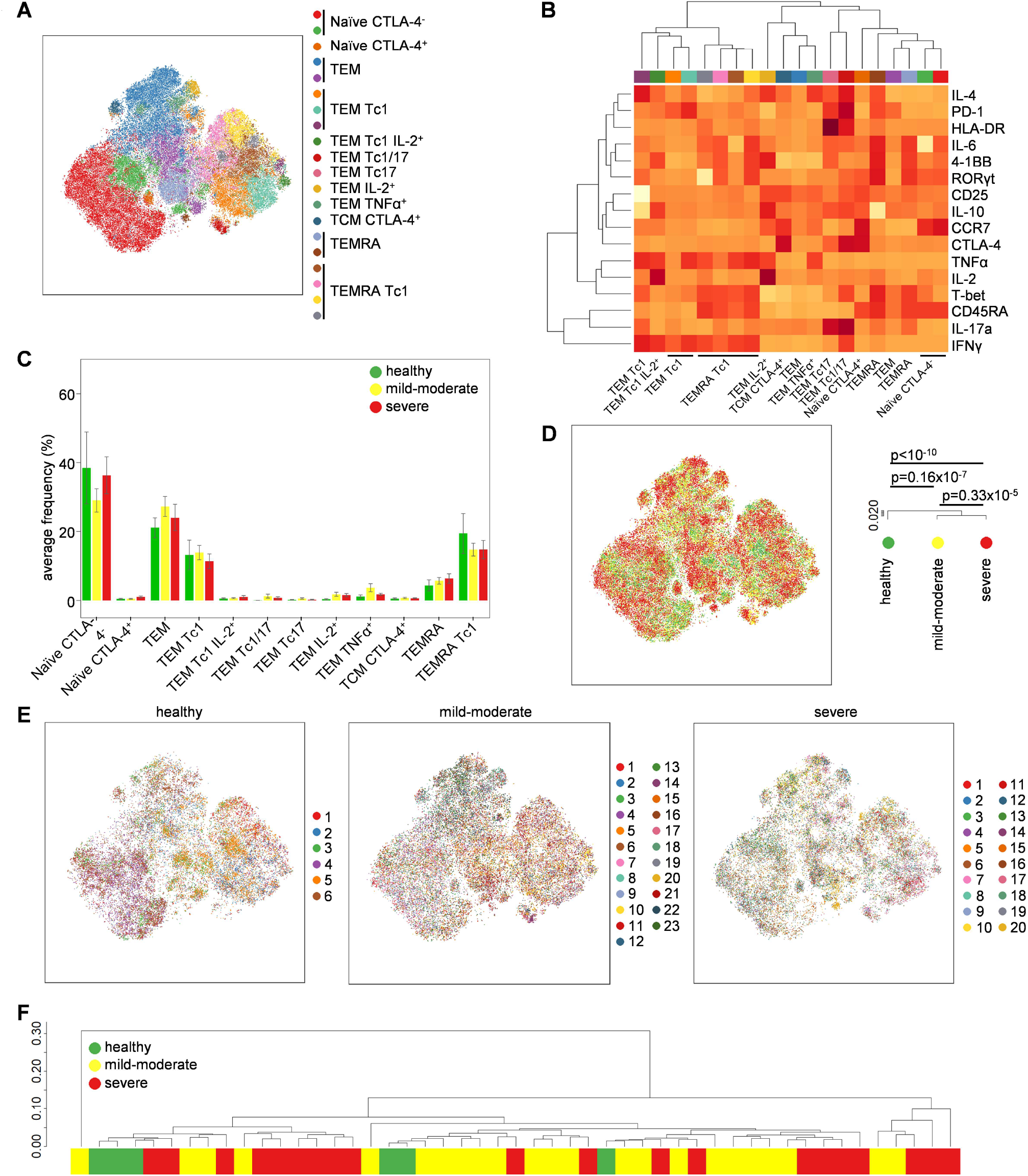
Profound activation of CD8^+^ T cells in both severe and moderate COVID-19. PBMCs were isolated from healthy controls (n=6), mild-moderate COVID-19 patients (n=23) and severe COVID-19 patients (n=20), stimulated and assessed through flow cytometry. CD3^+^CD14^−^CD4^−^CD8^+^ T cells were manually gated in FlowJo. **(A)** tSNE representation of CD8 T cell cluster populations based on the expression of phenotypic markers: CD45RA, CCR7, 4-1BB, CD25, CTLA-4, HLA-DR, IFNγ, IL-2, IL-4, IL-6, IL-10, IL-17a, PD-1, RORγt, T-BET, TNFα. FlowSOM clusters were annotated based on **(B)** expression of phenotypic markers across each cluster (see also Fig. S4). **(C)** Quantification of CD8 T cell subsets in healthy, moderate COVID-19 patients and severe COVID-19 patients. **(D)** tSNE representation of PBMC populations based on aforementioned phenotypic markers for each condition, namely healthy controls, mild-moderate COVID-19 and severe COVID-19 patients. The dendrogram shows the comparative similarity based on the Kolmogorov-Smirnov statistic calculated using the cross-entropy distributions derived from tSNE. **(E)** tSNE representation of PBMC populations based on aforementioned phenotypic markers for each condition, namely healthy controls, mild-moderate COVID-19 and severe COVID-19 patients, with each individual represented in a different color. **(F)** Dendrogram showing the comparative similarity between individuals based on the Kolmogorov-Smirnov statistic calculated using the cross-entropy distributions derived from tSNE. Mean ± SEM.

As a resource to the COVID-19 research community we provide in Supplementary Resource 1 the quantification of leukocyte subsets at an individual level, paired with demographic and clinical correlates. We further advocate for independent analysis of this resource, with the raw data available from https://flowrepository.org/experiments/2713.

## Discussion

In this study we present an in-depth investigation of the T cell compartment of mild to severe COVID-19 patients, using a platform with the capacity to investigate both intracellular transcription factor expression and, in parallel, functional cytokine production on a single cell level. Strikingly, COVID-19 patients that required hospitalization due to their condition harbored a peripheral T cell landscape that did not differ substantially from the healthy control one regarding the viral response. This is in stark contrast to the normal phenotype arising with viral infections, which specifically trigger Th1/Tc1-driven responses, with increased secretion of IFNγ and cytotoxic capacity. This condition has been particularly well studied in the case of influenza which could be considered to date one of the most prevalent respiratory viral infections ^40,41,42^ However, patients infected with SARS-CoV-2 do not display this distinct Th1/Tc1 polarization. While the mechanism for this polarization failure remains unknown, a potential explanation lies in the presence of a strong IL-6 environment ^9,43,44^ IL-6 is known to affect the Th1/Tc1 response by direct inhibition of IFNγ gene expression ^45,46^. Accordingly, this observation would support ongoing clinical trials testing the efficacy of tocilizumab in COVID-19 patients.

Our results are in line with recent studies regarding the absence of pro-inflammatory cytokines in the T cell compartment ^47^ or even a decrease of CD4^+^ secreting IFNγ^11^. These contrast with data identifying a Th1 signature in COVID-19 patients; the latter, however, being based on convalescent and non-hospitalized patients ^48^. Intriguingly, IFNγ levels in COVID-19 patient serum have been reported to be slightly elevated compared to healthy controls ^9^ If this is not due to differences in the patient cohort, the lack of a Tc1/Th1 phenotype would suggest a myeloid origin as the IFNγ source.

Our study was not able to unravel one of the most prominent features of COVID-19, the prevalent lymphopenia observed in most of the severely affected patients. In our study, both T and B lymphocyte compartments were affected equally, with a possible predominance for CD8^+^ T cells. Importantly, naïve T cells remained present in normal, or elevated, numbers, suggesting that T cell production/renewal was intact. Interestingly, this lymphocytopenia seems to be systemic, as previous studies in SARS-CoV-1 ^49^ and recently in COVID-19 patient autopsy revealed a pan-depletion in all secondary lymphoid organs, including satellite mediastinal lymph nodes with a disrupted architecture ^50^ (CONTAGIOUS manuscript in preparation). As massive lymphocyte infiltrates are not reported in lung anatomopathological investigations, lymphodepletion is unlikely to be explained by active recruitment of T cells to the lung tissues. An alternative cause of this lymphodepletion would be an increased T cell death, either through direct viral cytolysis or increased apoptosis through activation induced cell death (AICD). While lymphopenia may remain an epiphenomenon, the paucity of T cells may equally be contributing to disease. This is tentatively supported by results showing HIV positive patients with a slight trend toward poorer COVID-19 outcome ^51,52,53^. In addition, patients who received hematopoietic stem cell transplantation (HSCT), which leads to profound T cell lymphopenia ^54^, seem to have a worse outcome after SARS-CoV-2 infection, since preliminary data suggest 30% mortality according to an ongoing EBMT survey ^55^. On the other hand, the loss of B cells is less likely to contribute to disease, with patients who are genetically depleted of B cells showing normal outcomes ^56,57^.

While the overall T cell compartment did not display major differences in comparison to healthy controls, there were two particular enriched T cell subsets in more severely affected patients that could reflect the inflammatory condition present in COVID-19 patients. The inflammatory subset Tc1/Tc17 represents highly activated T cells with high expression of PD-1 and HLA-DR in addition to its ability to secrete both IFNγ and IL-17 cytokines. The presence of this population may reflect a deviation from the normal Tc1 anti-viral response caused by the pro-inflammatory IL-6-enriched environment ^58^. The contribution of this population to the inflammatory setting is, however, debatable, as the increase in relative number is mitigated by the lymphopenia, and no differences were observed between the mild-moderate and severe COVID-19 patients.

The only peripheral biomarker that did delineate disease severity was the increase in IL-10-producing regulatory T cells. Elevated IL-10 has been observed in the serum of COVID-19 patients before ^11^, however the cellular source was not elucidated. Production of IL-10 is a hallmark of activated regulatory T cells that reside in tissues such as the lung. This population, normally rare in healthy individuals, rose up to ~10% of the regulatory T cell pool in severe COVID-19 patients. The murine analogue to this population has a potent ability to limit inflammation and tissue damage triggered by microbial and environmental interactions at mucosal surfaces ^39,59^ In the case of viral infections of the lung, IL-10 restrains the development of IL-17-producing cells that damage the tissue ^60,61^, inhibits the innate inflammatory response to viral particles, and is likely beneficial in reducing the production of cytokines such as IL-6 that have been implicated in COVID-19 morbidity ^62,63^. Increase of this suppressive regulatory T cell subset could be a direct response to the progressing lung inflammation in COVID-19 patients, comprising a feedback inhibition circuit to prevent runaway inflammation and death ^64^ Potentially, elevated IL-10 could provide a blood-based biomarker for cases progressing to more severe lung damage. A more intriguing possibility is that individuals with higher IL-10-producing regulatory T cells exhibit defective adaptive immunity. IL-10^+^ regulatory T cells are symptomatic of many unresolved viral infections and are associated with long-term persistence. In respiratory infections, IL-10 potently suppresses anti-viral responses ^59,65^ and weakens the immune reaction to superinfection with bacteria ^66,67^ Since secondary infection leading to pneumonia is a major cause of death in influenza, and perhaps for some COVID-19 patients as well ^68,69,70^, excessive IL-10 production by regulatory T cells may be a key factor in COVID-19 outcomes. In principle, this truncated adaptive immune response could allow persistent infection and cause an over-reliance on innate responses, driving the pathological state. Under this latter model, early intervention (such as with IL-10 neutralization) could potentially restore appropriate adaptive immunity and quieten the excessively exuberant innate response in the tissue. However key replication, longitudinal and mechanistic studies would be first required, and IL-10 has proven stubbornly refractory to immune modulation in the past ^71^.

Key limitations in our study are the study size and the cross-sectional nature. The data presented here rely on a small number of patients without longitudinal follow-up, and lacking asymptomatic patients to screen the full spectrum of the COVID-19 immune signature. Study expansion and follow-up are ongoing to correct these deficits, and identify key components of the immune system responsible for a better protection against SARS-CoV-2. The other major limitation of the current work is the focus on T cells. With the limited phenotype changes observed here, a more comprehensive study of systemic myeloid cell changes in COVID-19 would be justified. Altogether, our study highlights the absence of a strong anti-viral response against SARS-CoV-2 across mild to severe COVID-19 patients, and the elevated presence of anti-inflammatory IL-10-producing regulatory T cells in the severely affected patients. This data suggests that a route to normalization of anti-SARS-CoV-2 immunity is critical in the attempt to cure these patients. Finally, with the intent to share and exchange knowledge to accelerate research on this pandemic, our data are available on the following open access repository for further investigation: https://flowrepository.org/experiments/2713.

## Methods

### Patient material

Healthy controls and symptomatic SARS-CoV-2-infected patients (qPCR confirmed) were collected as part of the CONTAGIOUS consortium (manuscript in preparation). Briefly, between March 27th and April 17th 2020, healthy volunteers and adult COVID-19 patients were recruited at COVID-19 hospitalization ward at UZ Leuven (Leuven, Belgium) after informed consent. COVID-19 diagnosis was made based on a positive qRT-PCR on respiratory sample and/or CT imaging compatible with SARS-CoV-2 disease. Patients were classified as mild-moderate (WHO clinical score 3-4) or severe (WHO clinical score 5-7) at point of sampling. All procedures were approved by the UZ Leuven Ethical Committee (protocol study number S63881). Blood samples from healthy controls and patients were stored at 15 °C for 3-6 hours prior to peripheral blood mononuclear cells (PBMCs) isolation using lymphocyte separation medium (LSM, MP Biomedicals) and freezing in liquid nitrogen. Whole blood count and differential values from the clinical laboratory were obtained concomitantly or within the 24 hours of the blood sample for the flow cytometry analysis.

### Flow cytometry

Frozen PBMCs were thawed, plated and incubated for 4 hours with complete RPMI containing phorbol myristate acetate (PMA 50 ng/mL), ionomycin (500 ng/mL) and Brefeldin A (8 μg/mL; all Tocris Bioscience) at 37 °C with 5% CO2. Cells were then washed twice with PBS (Fisher Scientific) and stained with live/dead marker (fixable viability dye eFluor780, eBioscience) and fluorochrome-conjugated antibodies against surface markers: anti-CD14 (TuK4), anti-CCR7 (G043H7) (eBioscience); anti-CD3 (REA613) (Miltenyi Biotec); anti-CD4 (SK3), anti-CD8 (SK1), anti-PD1 (EH12.1), anti-CD45RA (HI100) (all from BD Biosciences); anti-CD25 (BC96), anti-HLA-DR (L243), anti-CD40L (24-31), anti-4-1BB (4B4-1), anti-CD19 (HIB19) (all from BioLegend). Cells were fixed with 2% Formaldehyde (VWR chemicals) and then permeabilized with eBioscience permeabilization buffer according to manufacturer’s instructions. Cells were stained overnight at 4°C with anti-IFNγ (4S.B3), anti-IL-6 (MQ2-13A5), anti-IL17a (N49-653), anti-RORγt (Q21-559), anti-IL-2 (MQ1-17H12), anti-IL-10 (JES3-9D7), anti-T-bet (4B10), anti-CTLA-4 (BNI3), anti-GATA3 (L50-823) (all from BD Biosciences); anti-IL-4 (MP4-25D2), anti-TNFα (Mab11), anti-FOXP3 (206D) (all from BioLegend). Data were collected on BD Symphony (BD Biosciences). A maximum of 5×10^5^ events were acquired for each sample. tSNE, FlowSOM and heatmap analysis were performed in R (version 3.6.2). Raw.fcs data were compensated and dead cells and debris were gated out prior to file export. The complete set of FCS files used for the COVID-19 cytokine immune phenotyping has been deposited on FlowRepository and annotated in accordance with the MIFlowCyt standard. These files may be downloaded for further analysis from https://flowrepository.org/experiments/2713.

### Flow cytometry analysis

The concatenated dataset was analyzed through successive FlowSOM clustering and tSNE representation after exporting similar event numbers for each sample per condition group and then subsampling equal event numbers per condition. First, lineage markers (CD3, CD4, CD8, FOXP3, CD19, CD14) were used to separate leukocyte subsets. Second, activation markers (4-1BB, CCR7, CD25, CD45RA, CTLA-4, HLA-DR, IFNγ, IL-2, IL-4, IL-6, IL-10, IL-17a, PD-1, RORγt, T-bet, TNFα) were used to distinguish phenotypic clusters of each leukocyte subset, again using FlowSOM and tSNE. The characteristics of each identified cluster were assessed by means of histograms and heatmaps. Comparisons between groups (healthy, moderate COVID-19 and severe COVID-19) were performed with tests on the cross-entropy distributions of the tSNE representations of each group. In brief, for the original and t-SNE space of each tSNE plot, a probability per data point was calculated following the same approach as in the tSNE algorithm. From these point probabilities, the distribution of cross-entropy in the tSNE space relative to the original space was obtained for each group represented in the plot. All pair-wise comparisons between groups were evaluated with Kolmogorov-Smirnov tests on the difference between the cross-entropy distributions. Resulting p-values were corrected with the Holm method. Dendrograms were obtained from hierarchical clustering, using as distance the Kolmogorov-Smirnov statistic, that is, the L-infinity distance between the cross-entropy distributions (manuscript in preparation).

## Data Availability

All annotated primary flow cytometry data used in this study is openly available.

https://flowrepository.org/experiments/2713

## Acknowledgements

This work was supported by the VIB Grand Challenges Program, the KUL C1 program, the FWO Hercules program, the European Union’s Horizon 2020 research and innovation programme under grant agreement No 779295 (to A.L.), and the Biotechnology and Biological Sciences Research Council through Institute Strategic Program Grant funding BBS/E/B/000C0427 and BBS/E/B/000C0428, and the Biotechnology and Biological Sciences Research Council Core Capability Grant to the Babraham Institute. MG is supported by a fellowship from the Belgian Kid’s Fund. The authors acknowledge the important contributions of the COVID-19 clinical team and Pier-Andrée Penttila and the KUL FACS Core. We are grateful to Per Lungman who kindly provided us with data on COVID-19 disease after HSCT. The manuscript is dedicated to the memory of Michael Wakelam (Babraham Institute).

